# An attempt to optimize human resources allocation based on spatial diversity of COVID-19 cases in Poland

**DOI:** 10.1101/2020.10.14.20090985

**Authors:** Andrzej Jarynowski, Monika Wójta-Kempa, Łukasz Krzowski

**Affiliations:** Interdisciplinary Research Institute, Wroclaw, Poland; Department of Public Health, Faculty of Health Science, Wroclaw Medical University, Poland; Military University of Technology, Warsaw, Poland

**Keywords:** spatial modelling, optimal resources allocation, COVID-19, SARS-CoV-2

## Abstract

Our task is to examine the relationship between the SARS-CoV-2 arrival and the number of confirmed COVID-19 cases in the first wave (period from March 4 to May 22, 2020 (unofficial data)), and socio-economic variables at the powiat (county) level (NUTS-4) using simple statistical techniques such as data visualization, correlation analysis, spatial clustering and multiple linear regression. We showed that immigration and the logarithm of general mobility is the best predictor of SARS-CoV-2 arrival times, while emigration, industrialization and air quality explain the most of the size of the epidemic in poviats. On the other hand, infection dynamics is driven to a lesser extent by previously postulated variables such as population size and density, income or the size of the elderly population. Our analyses could support Polish authorities in preparation for the second wave of infections and optimal management of resources as we have provided a proposition of optimal distribution of human resources between poviats.

## Introduction

The optimal way to cope with SARS-CoV-2 global pandemic is still unknown, with different countries and regions having been more successful than others in controlling of SARS-CoV-2 spread (Jarynowski, Wójta-Kempa, Płatek, & Belik (2020); Jarynowski, Wójta-Kempa, Płatek, & Czopek (2020); Gelfand et al. (2020)). What has been acknowledged worldwide is that multi-dimensional approach must be applied, including medical, social and geographical factors. There is an urgent need to optimize the war with COVID-19 to reduce the danger of extension of epidemics and its consequences. Priority should be given to more efficient allocation of anti-epidemic resources which are already in possession, or which may be quickly and cheaply expanded in times of crisis (Trzos et al. (2017)).

The distribution of COVID-19 cases in Poland is very diverse and the reasons for this situation must be examined. In this article, however, we would like to use the already known and potential factors of COVID-19 spread to generate the model optimizing the allocation of human resources in order to defeat the plague. In spatial diversity of confirmed cases of COVID-19, some factors have been chosen to be examined in terms of their effect on infection dynamics. Demographic (like age, mobility, migration etc.), social (“income”,”PiS_support”) and COVID-related factors (population size,forest_density,population_density,arrival_SARS) are the ground for our proposal of proper sanitary staff allocation. Demographic factors, such as age structures, have shown a higher burden of symptomatic infections as well as mortality in countries with older versus younger populations (Jarynowski, Wójta-Kempa, Płatek, & Czopek (2020); Singh et al. (2020)), but regional distribution on the level of counties was not officially disseminated until summer 2020 (Raciborski et al. (2020)). We try to diagnose factors that catalyse and inhibit the dynamics of infections (possible causal relationship). Among others, we examine the hypothesis of the inequality paradox, through civilization backwardness (peripheral in the system of people flows (Bartosiak (2019))) and exclusion of selected social classes (e.g. elderly loneliness (Jarynowski, Wójta-Kempa, Płatek, & Czopek (2020); Duplaga (2020))) which affects a smaller fraction of elderly people in Poland in comparison with other EU countries (Raciborski et al. (2020); ECDC (2020)). Observations from US (Killeen et al. (2020)) and UK (Aldridge et al. (2020)) suggest that there are several socio-economic patterns such which vary substantially (in Poland too) and possibly increase the risk of infection, for instance relative poverty. Mobility, migration and, to a lesser extent, other demographic-social diversity patterns have already proved to be key factors in the dynamics of SARS-CoV-2 infections in a regional perspective, including USA (Messner & Payson (2020)), UK (Aldridge et al. (2020)), China (Jia et al. (2020)), Germany (Mense & Michelsen (2020); Felbermayr et al. (2020)), France (Pullano et al. (2020)), as well as internationally in Europe (Sannigrahi et al. (2020)) and around the World (Su et al. (2020)). Therefore, a similar picture could be expected also in Poland. Mobility and migration might play different roles in explaining variability of arrival time and number of cases, depending on the region of the country.

Many countries, especially low- and middle-income ones, experience many challenges when it comes to producing and customization of an adequate health workforce. Misallocation of health or sanitary workers, shortages of available health workers, and their limited productivity and performance may undermine efforts in restraining the disease. During a pandemic various sanitary inspections and their employees are a key link in slowing the disease spread. Sanitary Inspection employees (there is less than 10 000 sanitary workers in various inspections and hospitals in Poland overall, while in England - a country with a similar population - 18 000 new sanitary workers were hired during pandemic to work only with COVID-19 Mueller (2020); CNBC (2020)) have been extremely exhausted since March due to working on three shifts, also on Saturdays and Sundays. The main duties of Sanitary Inspection employees include epidemiological investigation, giving recommendations to local authorities, companies, etc. according to sanitary regime, isolating infected people, quarantining and surveillance of their contacts, writing dozens of reports, answering phone calls from residents, requesting and examining smear samples. In the dynamic pandemic situation there is an urgent need for a radical increase in employment in the Polish Sanitary Inspection - especially in State Poviat Sanitary and Epidemiological Stations.

In the COVID-19 pandemic, regional health authorities should decide how best to organize services in order to maximize the use of resources. The fundamental importance for efficient management in the region at the operational (poviat or voivodship) and tactical (outbreak or infected) level is mainly the possibility of optimal allocation of resources in time and space (Krzowski & Trzos (2020)), and this analysis is targeting this issue.

The aim of this paper is an exploratory and preliminary quantitative evaluation of the geographical spread on the level of county/poviat (NUTS-4) of SARS-CoV-2 virus (and COVID-19 disease caused by it) in Poland during the Spring wave of infections. There is a need for a spatial analysis of the spread of the SARS-CoV-2 virus in Poland in order to prepare detailed solutions in human resources area which would further result in taking actions minimizing the risk to public health during autumn/winter wave of infections.

### Epidemiology

Epidemiological data on the number of diagnosed cases in Poland in first 10 weeks after confirmation of first case on 04.03 formed epidemiological curve and indicate that the trend in the number of new diagnosed cases in most poviats in Poland was in a decreasing phase around 22.05. This means that the COVID-19 epidemic in Poland was already in a dying out phase outside of Silesia (Sitek et al. (2020)) and it continued plateauing until the end of summer for whole country. In 108 poviats, no new cases were recorded between 1-22.05.2020 (the reference elimination period of about 3 weeks). 6 poviats did not report even a single case before 22.05.2020. Unfortunately, official data on the incidence and testing of COVID-19 are only available with geographical accuracy to voivodship - corresponding to NUTS-3, so in our analysis we rely on data collected by volunteers in social surveillance project (Rogalski (2020)). However, it should be noted that the official data were provided by the Ministry of Health via Twitter in the form of an image and tables which were not easily accessible (Ochab-Marciniak (2020)).

There are plenty of forecasting models for Poland, for instance statistical (exmetrix (2020); StatSoft (2020); Chudy (2020); Śleszyński (2020)), or phenomenological ones (Krueger (2020b); Rakowski (2020); Gambin & Rosniska (2020); Mostowy (2020); Gonczarek & Wojcik (2020); Jasinski (2020); Jarynowski, Wójta-Kempa, Płatek, & Czopek (2020)). However, only a few models are targeting retrospective analysis of links between infection dynamics and explanatory variables such as population density (Sitek et al. (2020)), elderly population (Tarnowski (2020)), sex (Raciborski et al. (2020)) or tests numbers (StatSoft (2020)). Lessons from prior experience in various counties (poviats) should be drawn to assist the design of a future reaction.

### Data and methodology

The main purpose of a secondary analysis of existing registry data is to identify particular interactions of some economic and social variables with the dynamics of SARS-CoV-2 infections occurring in various areas of Poland. The level of analysis is the county/poviat (380 poviats) which corresponds to NUTS-4 level. The presented material is based on data obtained on the basis of:

1. Data on infections from sheets (Rogalski (2020)) based on poviat / voivodeship sanitary inspections and regional governments as well as journalistic investigations. The database with an accuracy to poviats is constantly improved and updated (in the article we present the data until 22.05), but it contains about 5% of missing data. Thanks to this database we can define the following explanatory variables:
  - arrival time (denoted as arrival_SARS) in days - we define it as the difference between the day of announcing the first confirmed case of the COVID-19 in a given poviat and 04/03/2020 - the first confirmed case in Poland. In the absence of positive cases in a given poviat, we assume a maximum value of 83 (we virtually infect these poviats on May 23, 2020);
  - Number of cases (denoted as size_COVID) - defined as the cumulative number of cases until 22/05/2020. There are available other non-official datasets with geographical distribution created by volunteers (UMW (2020); AmbasadaKultury (2020)), but they do not keep poviat NUTS-4 level as a reference. Due to unavailable data from some poviats, we have partly interpolated data from cities of Opole, Kielce, Radom, Koszalin, Siedlce, Ostrołęka, Płock, Piotrkow Trybunalski, Jelenie Gora, Skierniewice.
2. Registered data form GUS-Statistics Poland (GUS (2020)) - exact definitions are available from Statistics Poland:
  - earnings (Income) - normalized data on earnings in the economy in 2019 (100 - average for Poland);
  - afforestation (Forest_density) - % area of forests in 2018;
  - population_density - in person/km^2^ in 2019;
  - population_size - the number of people living in a given poviat in 2018 (normalized to maximum 100);
  - internal and external migrations (Internal / External _Emi / Immigration) - the number of people who change permanent residence for a given reason in 2017 for each poviat;
  - occupational structure (empl_ industry / agriculture / service) - number of persons working in the industrial / agricultural / service sectors in 2014, which can be linked with workplace-associated infections (GIS (2020); Krueger (2020a)) and level of remote working (Wojta-Kempa & Jarynowski (2020));
  - number of people in post-productive age >=65 in males and >=60 in females (postproduction_age) in 2019, as the number of older people has the potential to be associated with the size of the pandemic due to the greater vulnerability of the senior population to symptomatic disease and COVID-19 diagnosis (Jarynowski, Wójta-Kempa, Płatek, & Czopek (2020); ECDC (2020));
  - total sales of industrial enterprises (industry_revenue) in 2018 in million PLN. The data comes from various years due to the fact that after 2015 data on employment sectors are no longer provided on the poviat level. In order to analyse the interaction between variables, we have introduced mixed variables (imitating interactions between variables), e.g. the fraction of people in post-working age (senior_fraction) and the fraction of people employed in industry (industry_faction) to the entire population of the poviat.
3. Data from GUS Poznan on job mobility (GUSPoznan (2020)) from 2016-2017. Number of persons registered in the tax office covering municipality A (living) and working in municipality B (place of registration of the employer). The data has been aggregated to the poviat level. We built a directed weighted network of commuting patterns ^1^ :
  - the total number (mobility) of people entering and leaving a given poviat (this variable is not included in regression models due to co-linearity);
  - In-mobility (mob_in) - the sum of weights of incoming edges (the number of people working in a given district and living in another) of the mobility graph;
  - Out-mobility (mob_out) - the sum of the outbound edge weights (the number of people residing in a given poviat and working in another) of the mobility graph;
  - Pagerank, Closeness, Betweenness centralities are derivative centrality functions (Jarynowski, Paradowski, & Buda (2019));
  - Logarithm with a base 10 of total mobility (log_mobilty).
4. Geographic coordinates of poviats’ centroids (longitude / latitude) in degrees. We also calculated the radial distances between centroids of poviats d(x, y) in degrees. We have created measures of geographical dependence for each poviat y with the influence of other 379 poviats x as:

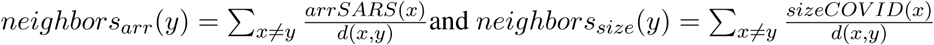
5. Results from the National Electoral Commission (PKW (2020)) in European Parliamentary election - support for PiS (Law and Justice) ruling party. The dataset was chosen because of the fact that due to the lack of data from surveys with poviat accuracy, election preferences could be good indicators of social attitudes in local communities (Jarynowski & Klis (2012)).
6. Air Quality (GIOS (2020)) interpolation for each poviat. Mean particulate matter: PM_10_(denoted as PM) concentration in *μ* g/m^3^ over the years 2012-2015.

All explanatory and explained (sizeCOVID(x), arrSARS(x)) variables are numeric and satisfy ratio scale condition. The main statistical approach is calculating multiple regressions with Akaike selection criteria on the SARS-CoV-2 arrival time to each poviat and the number of COVID-19 cases based on socio-economic variables. Multidimensional scaling techniques were also applied. It should be noted that PCA clustering differs from hierarchical clustering [Fig. 1], for instance in including also significant opposite relations into one cluster. Moran’s spatial autocorrelation was applied to measure similarity in infection dynamics between nearby poviats.

**Figure 1.**
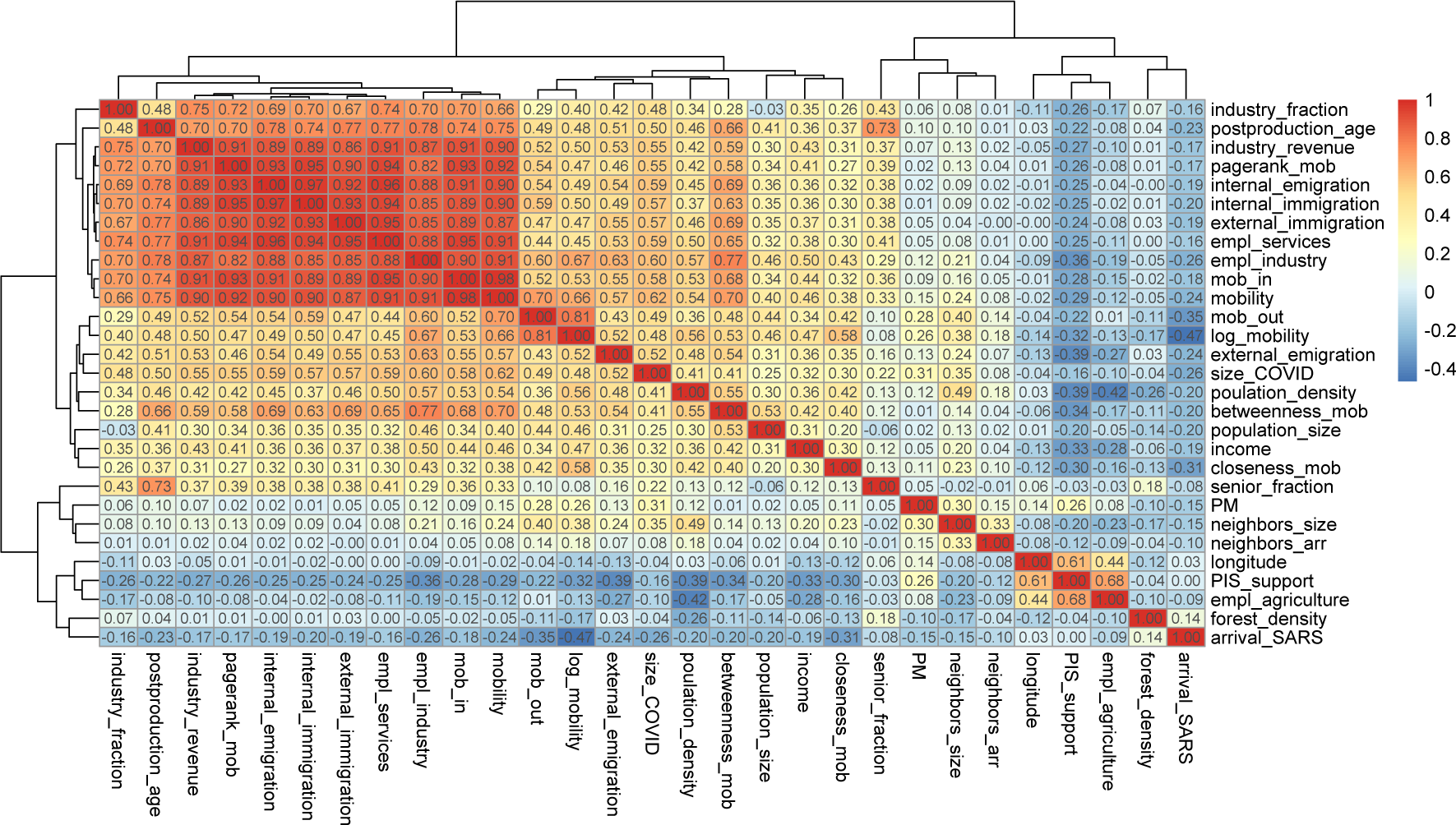
Pearson correlation matrix between variables taken into account in the hierarchical system analysis

## Results

### Correlations, regressions and multidimensional scaling

There is a significant difference between disease dynamics in Silesia and the rest of Poland (Śleszyński (2020)) and our task is to present this complex structure. The most significant factor for the progression / propagation of the infectious diseases is often human mobility (Belik et al. (2011)). In the hierarchical system (dendrograms) of correlation matrix, one can notice a cluster of strongly positively correlated variables such as simple mobilities, age and industrialization (top left Fig. 1). The next cluster contains variables related to population, complex mobility measures and the number of COVID-19 cases (middle bands Fig. 1). The last cluster (bottom right Fig. 1) refers to variables slightly negatively correlated which are related to geographical and social aspects as well as SARS-CoV-2 arrival time.

We have calculated Moran’s I - measures of spatial autocorrelation with inverse radial distance (1/d(x,y)). We proved that arrival time is slightly and outbreak size is moderately related with the locations where they were measured (both correlations are statistically significant) [Tab. 1].

**Table 1:** Moran’s I - spatial autocorrelation for arrival time and outbreak size for each poviat.

|  |  |  |  |
| --- | --- | --- | --- |
| Moran's I | arrival time | No. cases | expected |
| estimate | 0.0234 | 0.084 | -0.003 |
| p-Value (is the difference between observed and expected significant?) | <0.001 | <0.001 | NA |

Linear regression of the full model [Fig. 2] indicates a small (on the verge of statistical significance) role of geographical proximity of the appearance of the first cases for arrival time to a new place and a small (but statistically significant) role of clustering poviats with a similar number of cases.

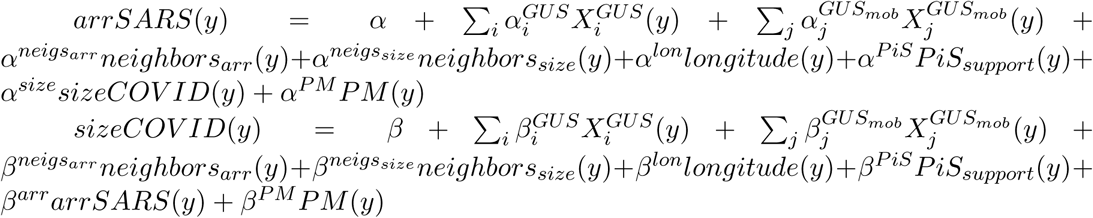

Where parameters for both depending variable *β, α* are intercepts (without indexes) and fitted parameters (with indexes), *X*^*GUS*^ and *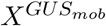* are variables described in chapter Data and Methodology in points 2 and 3 respectively.

**Figure 2.**
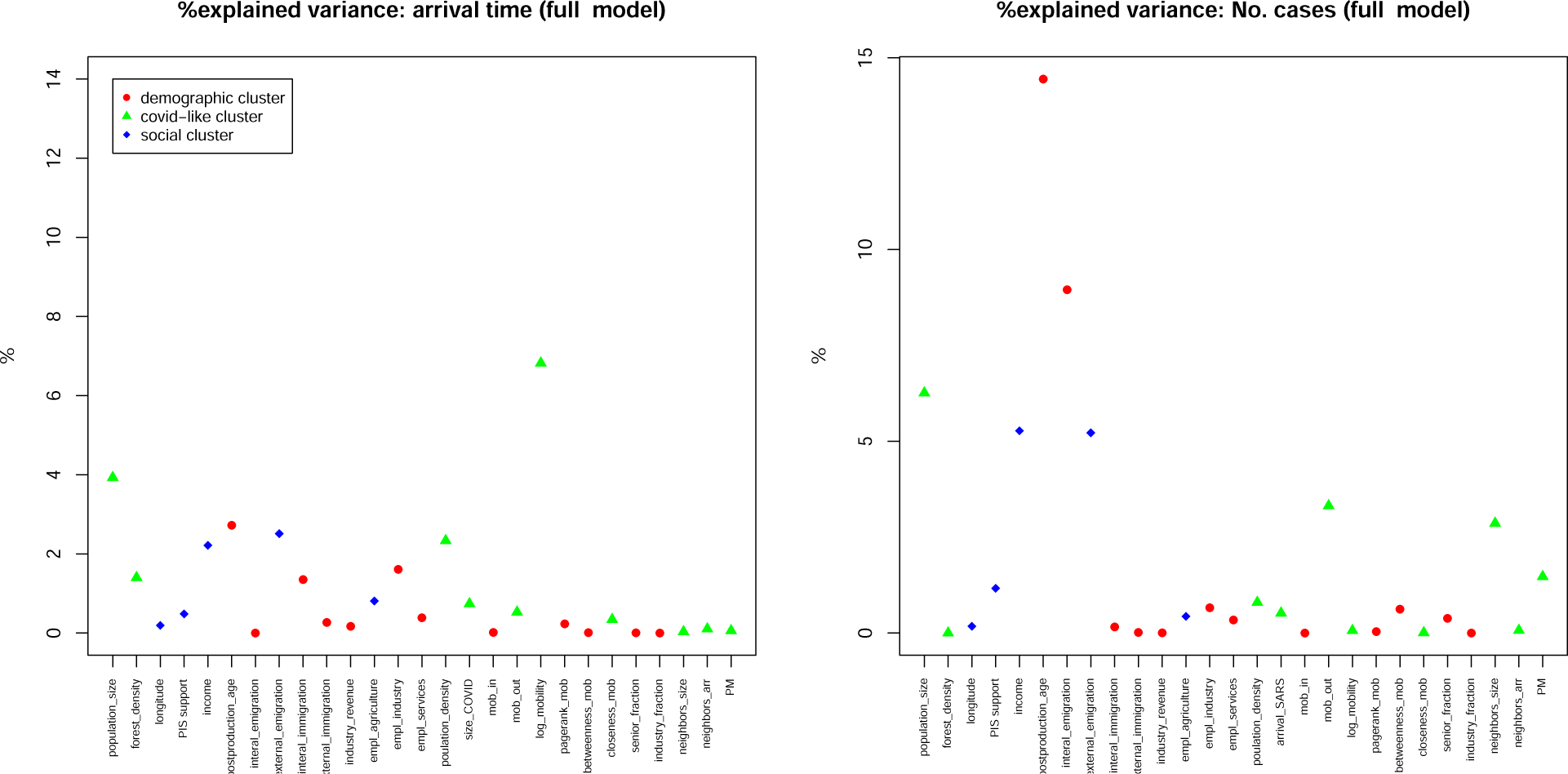
Predictive model (Multiple linear regression) of arrival times (left - the model gives 29% explained variance) and number of cases (right - the model gives 53% explained variance) on all non-collinear (linear combinations of other variables) variables. The y-axis shows the % of explained variance for each of the variables from the x-axis. The points are marked and colored according to the PCA classification (as in the same single legend for both charts)

The arrival time in poviats could be explained (6%) by logarized mobility, which is evidence of the non-linear nature of relationships. The number of cases could be explained (14%) by the number of people in post-productive age. This is certainly associated with over-representation of the symptomatic course of SARS-CoV-2 infection within the elderly (ECDC (2020)), but socio-economic relationships are also possible. The next factors are emigration: internal (9%) and external (5%) and outgoing mobility (4%), which may be associated with a wave of returns from the area of high SARS-CoV-2 prevalence, possibly also influenced by reasons of socio-economic nature. Other important factors are also the population size (6%) and income (4%).

Principal component analysis (PCA) on all variables reveals 3 main clusters also marked in Fig. 2 (explained variance in bracket):

- demographic (45%) - (“postproduction_age”, “internal_emigration”, “internal_immigration”, “external_immigration”, “industry_revenue”, “empl_industry”, “empl_services”, “mob_in”, “mobility”, “pagerank_mob”, “betweenness_mob”, “senior_fraction”, “industry_fraction”) taking into account variables related to the elderly, employment structure and mobility;
- covid-like (11%) - (“population_size”, “forest_density”, “population_density”, “arrival_SARS”, “size_COVID”, “mob_out”, “log_mobility”, “closeness_mob”, “neighbors_size”, “neighbors_arr”, ‘PM’) taking into account variables related to pandemic, population size and density, air quality, spatial correlates and non-linear migration;
- social (8%) - (“longitude”, “PiS_support”, “income”, “external_emigration”, “empl_agriculture”) taking into account variables related to distance to Germany, income, emigration, agriculture and social attitudes linked to voting preferences.

### Selective multiple regression

The transmission dynamics of infections in the already affected areas is mainly determined by behavioural factors such as the structure of contacts and compliance with risk mitigation recommendations (Jarynowski, Wójta-Kempa, Płatek, & Czopek (2020); Jarynowski, Wójta-Kempa, Płatek, & Belik (2020)). Structure of transmission routes in Poland (GIS (2020); Krueger (2020a)) and worldwide (ECDC (2020)) suggests that the main transmission route are intensive and longterm contacts (home and occupational infections have about 75% share of infections). Mobility/Migration has a different function in catalysing transmission in an already infected area. Based on analyses among smartphone users with activated geolocation and Internet connection, the decrease in mobility to baseline values during the most extensive restrictions in the general population was in the range of 30-70%(Krueger (2020b)), and mobility has slowly returned to normal levels since restrictions were lifted (Krueger (2020a)). It is worth to mention that mobility in Silesia decreased to a significantly lesser extent during the restriction period than in other areas of Poland (Selectivv (2020)). The main model was selected (without taking into account spatial correlations and interactions between the number of cases and the arrival time to show explanatory variables only) by the mixed backward / forward selection method using the Akaike criterion [Tab. 2]. We chose linear regression as a zero approach, while each of our depending variables could be adopted by separate model. Starting point includes:

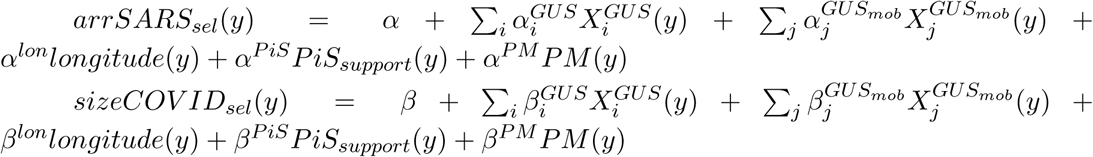

and the final models are composed with selected (most important) variables in Tab. 2. Selected regressions [Tab. 2] need to be interpreted together with correlation matrix [Fig. 1] and full model [Fig. 2], because we are dealing with a strongly correlated system.

Other network centralities (Jarynowski et al. (2014)) based on employee flow networks, i.e. closeness, pagerank and betweenness, also correlate with explained variables (particularly, closeness correlates with the time of arrival and pagerank with the number of cases). Due to the observed non-linearities, the logarithm of mobility was also taken into account and it strongly correlates with the arrival time. After mobility / migration, the employment in industry correlates the most with the number of cases [Fig. 1]. Due to population flows in the form of commuting and interaction between employees, industry is a sector that is impossible to be digitalized and switched to remote work (PolskiInstytutEkonomiczny (2020)). The arrival time correlates the strongest with mobility / migration (especially the outgoing direction, which may suggest above all the outflow of employees, especially abroad and could be associated with their returns to Poland). Professional activity was also important for the rapid emergence of the virus, especially in the field of industry.

Strong effect sizes of mobility in all dimensions and structure of employment/industrialization in the selected model for number of cases (fit R^2^∼0.5) shows the socio-economic conditioning of a epidemic [Tab. 2]. Regions with a highest employment rates (mainly in poorly paid industry and service) have significantly higher burden of COVID-19. PM_10_ is also a statistically significant catalyst of the outbreak size, hence it can be said that air quality could moderate susceptibility to SARS-CoV-2 infection Conticini et al. (2020). Poor-quality air has been already known to be a risk factor contributing to the development of respiratory diseases (Kowalski & Konior (2020)). The fit of the selected model for arrival times is on the level of R^2^∼0.3 only and mobility, internal immigration and international emigration, support for the PiS (ruling) party (which is indirectly associated with the social structure of the electorate (Jarynowski & Klis (2012))) significantly accelerated the appearance of the first SARS-CoV-2 cases. In addition, a longitude (proxy of distances from economic attractors in Western Europe, in which SARS-Cov-2 arrived earlier than in Poland) is borderline significant. It is worth noting that the size of population, density (which was a contradiction to correlation and non-selective multiple regression model) as well as income of individuals do not significantly differentiate the infection dynamics in a selective model [Tab. 2].

### Optimal resource allocation

Taking into account the spatial data-driven methodology, we would like to consider the distribution of needs for empowering and extending public health/sanitation workers. To choose the most vulnerable counties to be supported with the highest priority, we propose combination of criteria obtained from selected model [Tab. 2]:

- high predicted No. cases (to support poviats with the highest level of prognosed cases);
- high Residuals in No. cases (to support poviats which have more cases than it’s predicted by the model, because our model was trained only on limited predictor space and maybe there are other non-random important factors);
- short predicted arrival times (to support poviats which are the most likely to be the earliest to have new wave of cases).

We have standardized each criteria and proposed a composition of a zero approach optimization schema:

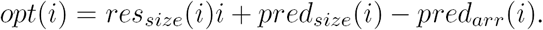

where *i* is a given poviat, *opt* is optimization goal function, *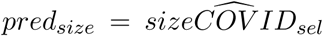* is predicted value of outbreak size from linear model with parameter estimation from Table 2, 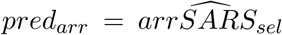 is predicted value of arrival from linear model with parameter estimation from Table 2, *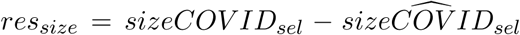* residuals is difference between actual outbreak size and prediction of the model for outbreak size.

**Table 2:** Multiple linear regression (selection by Akaike criterion). Dependent variables: arrival times and number of cases (NOTE - other selection models and other missing data interpolation schema and in-depth analysis of the model are described in the codes)

| Regression for each Polish poviats till 23.05 | Arrival time (in days) of first SARS-CoV-2 case in poviats |  | No. confirmed SARS-CoV-2 cases in poviats |  |
| --- | --- | --- | --- | --- |
|  | Estimates | p | Estimates | p |
| (Intercept) | 98.2081 | **<0.001** | -128.7163 | **<0.001** |
| forest_density | 8.0467 | 0.134 |  |  |
| longitude | 0.6048 | 0.12 |  |  |
| PIS support | -0.209 | **0.026** |  |  |
| postproduction_age | -0.0001 | 0.096 |  |  |
| internal_emigration | 0.0026 | 0.111 |  |  |
| external_emigration | -0.0118 | 0.051 | 0.1443 | **<0.001** |
| internal_immigrat | -0.0024 | **0.043** |  |  |
| empl_agriculture | -0.0003 | 0.149 |  |  |
| mob_in | 0.0002 | 0.076 |  |  |
| mob_out | 0.0005 | **0.042** | 0.0043 | **<0.001** |
| log_mobility | -8.6407 | **<0.001** |  |  |
| income |  |  | 0.5229 | 0.158 |
| industry_revenue |  |  | -0.0026 | **0.034** |
| empl_services |  |  | 0.0015 | **<0.001** |
| betweenness_mob |  |  | -0.0052 | 0.051 |
| PM |  |  | 2.6145 | **<0.001** |
| Observations | 380 |  | 380 |  |
| R<sup>2</sup> / R<sup>2</sup> adjusted | 0.279 / 0.257 |  | 0.496 / 0.486 |  |

Applied optimization schema is supposed to favour No. cases (2 normalized variables) over arrival time (1 normalized variable). Let us consider allocating 1000 new Disease Intervention Specialists for a better outbreak investigation and providing communication capacity of already overloaded channels. We decide to support counties only with positive opt proportionally to its value to get first attempt to the optimal allocation [Fig. 3].

**Figure 3.**
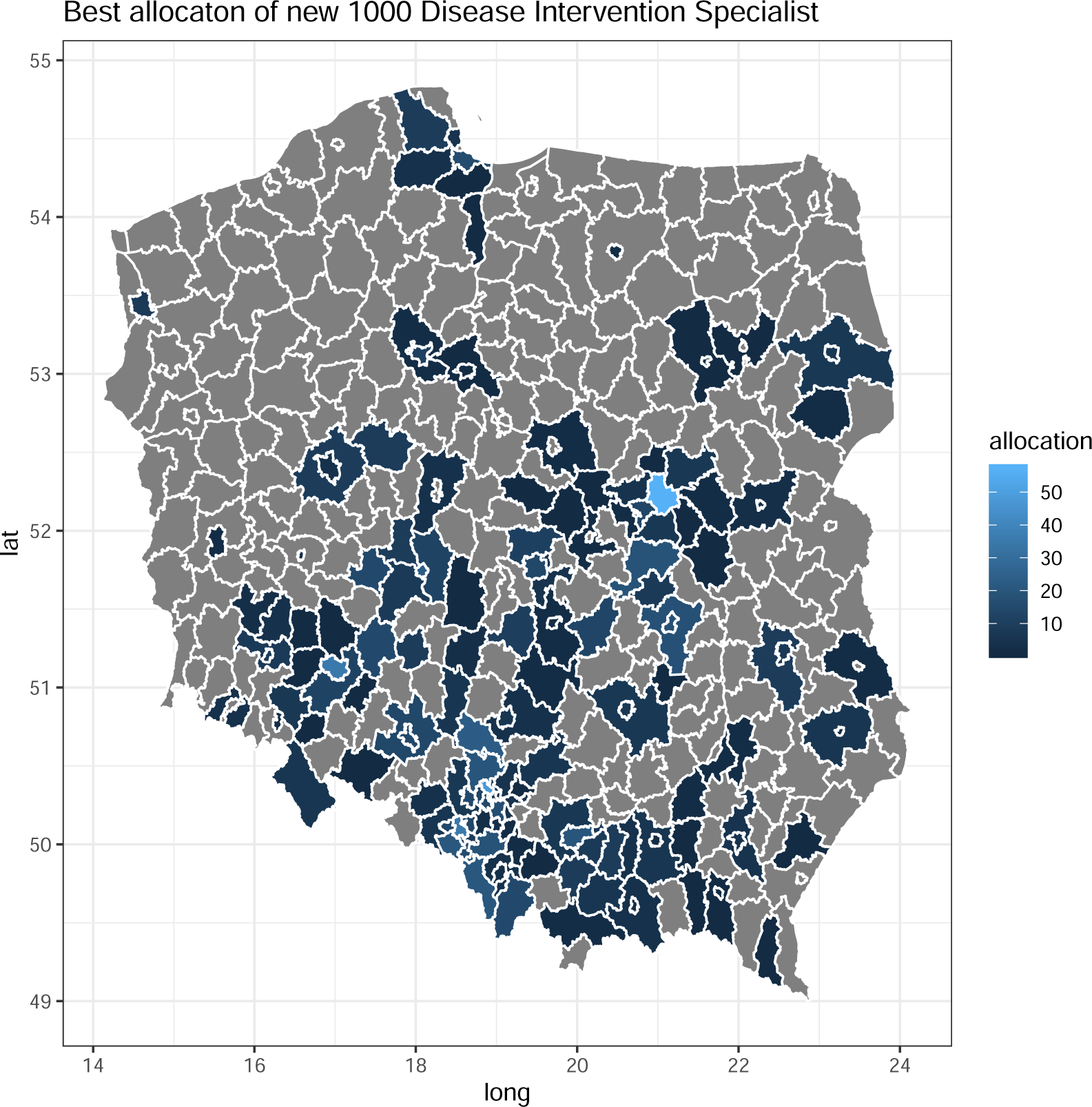
Optimal spatial allocation of 1000 new Disease Intervention Specialists

This example illustrates a zero approach, which counties would need additional staffing in a simple form of optimization function taking into account equally predicted No. cases, Residuals in No. case and arrival times. Further optimization analysis should be conducted, however we could recommend to think about supporting poviats as shown [Fig. 3].

Considering proper allocation of sanitary workers, we assume that it should overlap with the actual path of spreading the infections. On the map we can see several strategic regions (poviats) which should be empowered by new sanitary workers. In the presented illustration, we present the option of allocating new employees of sanitary services. 40-50 new employees should be located in the most risky zones to support the fight against the raging epidemic. These zones are Warsaw and Wroclaw, Krakow, Silesian Agglomeration, but also other large cities with high population density (Tri-City). Regions requiring additional investment are also poviats from the Opolskie, MaÅopolskie and Lodzkie voivodships. The Authors are aware, that second wave of Covid-19 in Autumn may bring another local epidemic outbreaks. The presented map is the visualisation of possible action, based on spatial numbers of infections. Its main aim is take first step to discuss the optimal way to cope with infectious threat.

## Conclusions

Our spatial analysis of disease incidence can be an example of the application of social geography in public health. Since August, mitigation strategies in Poland have been regionalised on the poviat level. The analysis of poviat characteristics in the context of the number of cases of disease is not only an attempt to understand the current situation, but also to initiate a discussion on the optimal distribution of resources necessary to fight health threats, such as epidemics of infectious diseases.

The most important conclusions carried out from the analyzes are:

1. SARS-CoV-2 infections classified at the poviat level are detected in the wide belt in the Southern part of Poland. In the analyzed period, the highest number of infections occurred in the following voivodships: Slaskie, Mazowieckie, Malopolskie, Wielkopolskie and Dolnoslaskie. COVID-19 hit the most early and strongly the most connected and richest regions (e.g. the Silesian agglomeration, Warsaw, Wrocław), and the least those which are poor and less-connected (e.g. in the belt of relative poverty from Lubuskie to Mazury (Śleszyński (2018))). The pandemic reached the economic periphery statistically significantly later (comparing arrival times) and to a lesser extent (comparing the number of cases) [Fig. 1, Tab. 2]. However, the outflow nature of regions with a large number of Polish emigrants significantly accelerated the appearance of the first cases and outbreaks there. Imported cases were the most frequent category of confirmed cases for the first 3 weeks after official introduction to Poland (Krueger (2020a); Jarynowski, Wojta-Kempa, et al. (2020)). SARS-CoV-2 virus officially arrived later in less urbanized (e.g. heavily forested) areas.
2. The analysis showed that some features of poviats (administrative units) are conducive to disease. Poviats with large workplaces employing a lot of stationary commuters (“low-wage industries”) became large hotspots during the first wave of the epidemic. The poviats in the western belt, with the predominance of smaller establishments or workplaces enabling remote work, did not record so much outliers from the average number of cases.
3. The impact (especially negative) of social inequalities was visible, e.g. in the US and UK, where poorer counties (with a large fraction of the poor) were significantly less able to combat COVID-19 (Gelfand et al. (2020); Messner & Payson (2020); Bartscher et al. (2020)). This is also in agreement with other European Union studies, where regions with high social capital are more prone to high COVID-19 notification numbers, especially in the first phase of the outbreak (Bartscher et al.(2020)). However, this is not a case in Poland (income is a catalyst of infection dynamics, although not a significant one [Fig.1, 2]). The hypothesis about perpetuating inequalities (Domanski (2020)) is unlikely to work out on an ecological level (aggregated to the poviat). Also data on population mortality in individual terms from Europe (e.g. from Sweden) suggest that income could increase (in some cohorts) the risk of death from COVID-19 (Drefahl et al. (2020)).
4. Importantly, the likelihood of mass tourism and business travel had no effect on explaining the variability of either the number of cases or the arrival times, which distinguishes Poland from other countries like Germany (Felbermayr et al. (2020); Kovacs et al. (2020)) or USA (Killeen et al. (2020)). This can be explained by the still low purchasing power of Poles (an average Pole travels abroad touristically 5 times less often than a German (Delhey et al. (2019))). The lack of significance of external immigration could be associated with its marginal size in 2017 (Statistics Poland does not provide newer data), and perhaps with current data the effect of the inflow of foreign immigrants (Górny & Śleszyński (2019)) would be more pronounced due to the exponential increase of immigrants in the last few years e.g. in Greater Poland (Paradowski et al. (2020)). Immigration will not affect the spread of SARS-CoV-2 in Poland in such an extend during Autumn/Winter wave due to reduced international mobility and obligatory quarantine for travelers outside of EU.
5. Global centrality measures (such as closeness, pagerank or betweenness) do not explain the variability in infection dynamics better than local measures (total weighted as well as incoming and outgoing vertex centrality), in which Poland is also different from other countries (Jia et al.(2020); Maier & Brockmann (2020); Felbermayr et al. (2020); Pullano et al. (2020)). This could be explained by relatively short chains of infections, which is probably associated with the simultaneous introduction of the virus into a large number of poviats (Pybus (2020)), which were the source for only one neighbouring poviat on average. This interpretation could be also confirmed by the fact that the geographical proximity of already infected poviats did not significantly affect arrival times in terms of regressions [Fig. 2] or in much smaller extent to number of cases in Moran’s correlations [Tab. 1]. Only the number of cases in neighbouring poviats significantly affects the arrival time and the number of cases in a given poviat.
6. The elderly population in Poland has relative small impact on infections dynamics (especially nursing homes and long term care facilities, which are very often picked by media, but they comprise only 3% of all diagnosed cases from “Epibaza/SRWE dataset” (Krueger (2020a))). However, these numbers must be interpreted with caution, because in “National Institute of Cardiology dataset” especially nursing homes and long term care facilities account for 13% infections in a similar time frame (Raciborski et al. (2020)). The size or fraction of the senior population is losing its importance in the selected model [Tab. 2].
7. Population density or population size lose predictive power against the infection dynamics in favour of other variables too [Tab. 2]. This may mean that the size of the pandemic is primarily related to the socio-economic conditioning of the regions and in a much smaller extent to demographics such as age structure or population density.
8. A statistically significant relationship between the number of cases and the support of the currently ruling party reveals the likely relationship between PiS electorate (large industrial and mining plants and medium-sized cities, a large percentage of older people, high level of emigration etc.) and vulnerability to outbreaks. It may also be related to the level of effectiveness of selected in local elections authorities in managing the epidemic at the poviat level. Moreover, areas that have proven experience in the fight against infectious diseases (e.g. nosocomial infections as NDM-2 stains Jarynowski, Grabowski, et al. (2019)) such as Poznan proceed better than it would be predicted by a simple socio-economic model classification.
9. The additional recruitment of sanitary services during the second wave of Covid-19 in Poland is one of the priorities of the epidemic management. Our proposal to allocate new sanitary workers is based primarily on a spatial analysis of the number of infections. In our opinion, the proper allocation of human resources can contribute to the launch of quick paths to react to new outbreaks, especially where stationary work is the basis for the implementation of tasks (production, processing plants, etc.).

To sum up, mobility/migration and occupation structure in geographic regions associated mainly with industry and employment rates play the most important role in infection dynamics Poland. However, this paper is just an ecological (on the level of poviat) retrospective correlation study and its low position in hierarchy of evidence-based medicine must be stressed. It has also a plenty of limitations, such as controlling only for selected variables. For instance, there are definitely differences in testing strategies (not considered in our analysis), for example 20% of share in total number of test are coming from Warsaw region, which has only 5% share of total population (Rogalski (2020)). The greatest bias factor is the unofficial character of surveillance, as it led, for instance, to the fact that missing data in infection dynamics caused interpolation modification which was applied for almost 5% of poviats. Statements similar to those presented in this article can also be applied to the number of deaths classified due to Covid-19. Their overtones would perhaps be a stronger argument for the central authorities.

## Discussion and Recommendation

Lockdown (closing schools and most nonessential businesses) was proved to be the most effective intervention slowing down COVID-19 epidemic (Haug et al. (2020)). However, it does not seem to be cost-effective. Spring lockdown costs in UK (Miles et al. (2020)) was roughly estimated to 300k GBP/QALY, which exceeded 10 holds profitable threshold. In Poland, the restrictions and measures already applied during Spring cost the economy almost 50 billion EUR (Pinkas et al. (2020); Raciborski et al. (2020); Ministerstwo_*F*_*inansow*_*P*_*l* (2020)), with roughly estimate ∼500k PLN/QALY, which exceeded also 10 holds profitable threshold (Infodemia_*P*_*olsce* (2020)). Thus, the most cost-effective (except Standard Precaution (Rosiński et al. (2019)) and proper communication (Jarynowski, Wójta-Kempa, & Belik (2020a,b))) mitigation technique: “trace, track, treat and isolate” could be applied during the second wave to a much greater extent and with geographical precision as we proposed in this paper.

Healthcare systems must be accessible, effective, and adequately respond to unexpected situations such as an outbreak of an infectious disease. In Poland, healthcare systems are heavily burdened with patient demographics (mainly due to age) and suffer from labor shortages. That is why it is so important to make the right decisions in relation to the current situation, so as not to increase inequalities in access to healthcare. Providing adequate access to medical personnel in the country (especially at the place of residence) based on the needs of its inhabitants is one of the most important challenges of modern health policy. However, the most satisfactory approach is to plan the optimal human resource allocation, in 2020 we have to act quickly to minimize the consequences of SARS-CoV-2 epidemic. Epidemiological services (especially the Sanitary Inspection) should be particularly strengthened in highly connected regions and with industrial / high employment characteristics, so that they could quickly and efficiently investigate and close outbreaks (Krueger (2020b)) in workplaces. Certainly, Sanitary Inspection should keep monitoring and investigating outbreaks in houses, nursing homes (because of high case fatality rates there), hospitals, schools etc. as before, yet workplaces should be prioritized. At the same time it should be noted that for many Regional Sanitary Inspection units the budget planned for the entire 2020 was exceeded already in April (WSSElubuskie (2020)), and central/local governments in the regions co-finance them to varying degrees. Identifying particularly endangered poviats is intended to support the fight against the pandemic (especially staffing for the purpose of in-depth and rapid investigation of infection outbreaks) and under no circumstances can it be interpreted as stigmatizing individual poviats and their residents, which is associated with detached risk perception (Jarynowski, Wójta-Kempa, & Belik (2020b)).

Proper data is a key to understand and prepare for Autumn/Winter wave of infections (Duszynski (2020)). Moreover, we regret that most of Polish researchers cannot work of official detailed surveillance data, while data used by institutions within Ministry of Health seems to be at least inconstant in comparison with public available resource.

We recommend that during the next wave of infections, Polish Sanitary Inspection should be strengthened in terms of staff and IT services in high risk poviats.

## Data Availability

Data and codes available: https://github.com/ajarynowski/Spatial_Covid_Poland

## Acknowledgements

Data and analyses have been deposited in an interactive simplified form: https://www.kaggle.com/andrzejjarynowski/spatial-covid-19-in-poland full R Markdown and data https://github.com/ajarynowski/Spatial_Covid_Poland and project description http://www.infodemia-koronawirusa.pl/geografia-zakazen-w-polsce. Data availability is a crucial element of COVID-19 epidemiology Szmuda et al. (2020), so we hope this dataset will be of use to other researchers, mainly Polish students. The authors would like to thank the European Commission (EOSC-48) for partial support of data collection, the NCN (2016/22/E/HS2/00034) and Karolina Czopek for proofreading, epidemiologists Vitaly Belik, Ireneusz Skawina, economists Mariola Chrzanowska, Andrzej Buda and sociologist Daniel Płatek geographer Przemysław Śleszyński for consultations.

## Footnotes

1 It is worth to mention that commuting patters could be different: some of people are commuting on the daily basis, some returning home for weekend, others only for holidays, etc.

